# Comparative Effectiveness of Rituximab and Cladribine in Relapsing-Remitting Multiple Sclerosis: A Target Trial Emulation

**DOI:** 10.1101/2024.12.10.24318773

**Authors:** Brit Ellen Rød, Einar A. Høgestøl, Øivind Torkildsen, Kjetil Bjørnevik, Jon Michael Gran, Mathias H. Øverås, Marton König, Elisabeth G. Celius, Kjell-Morten Myhr, Stig Wergeland, Gro O. Nygaard

## Abstract

**Background:** Head-to-head comparisons of high-efficacy therapies for relapsing-remitting multiple sclerosis (RRMS) are lacking. We emulated a target trial to compare the long-term effectiveness of rituximab and cladribine.

**Methods:** We estimated the effect of initiating treatment with rituximab or cladribine by emulating a target trial using data from the Norwegian MS Registry and Biobank at two university hospitals with different treatment strategies. Cumulative incidence and risk differences after 4 years were estimated using a weighted Kaplan-Meier estimator, adjusted for baseline covariates. The primary outcome was MRI disease activity, with the secondary outcomes including relapses and safety.

**Results:** The study included 285 patients, 159 receiving rituximab and 126 receiving cladribine, with a median follow-up of 4.5 years (IQR 4.0 to 5.0). Rituximab-treated patients had a lower risk of new MRI disease activity compared to cladribine-treated patients (*p* < 0.0001). The 4-year risk was 18% (95% CI 11 to 23) for the rituximab-treated patients and 57% (95% CI 48 to 65) for cladribine-treated patients, yielding a risk difference (RD) of 38 percentage-points (95% CI 29 to 51). The 4-year RD for relapse was 11.2 percentage-points (95% CI 3 to 18) and the RD for discontinuation or a third dose of cladribine was 13.7 percentage-points (95% CI 9 to 25). The incidence of hospitalizations related to potential adverse events was 6.0 per 100 person-years for rituximab and 4.1 per 100 person-years for cladribine.

**Conclusions:** These findings suggest that rituximab has superior effectiveness compared to cladribine during a median follow-up of 4.5 years.

## INTRODUCTION

High-efficacy therapies have improved clinical and radiological outcomes for patients with relapsing-remitting multiple sclerosis (RRMS)^1^. Despite their extensive use^2^, head-to-head randomized controlled trials (RCTs) are lacking, leaving the question of which therapy is most effective unanswered.

Rituximab, an anti-CD20-antibody used off-label, and cladribine, a purine analog, have been used extensively in Norway as first- and second line treatments for RRMS. Randomized trials have demonstrated their high efficacy in reducing magnetic resonance imaging (MRI) disease activity and improving clinical outcomes^3–5^. However, the lack of comparative studies has led to variations in treatment preferences across centers, influenced by factors such as physician preferences, hospital policies on off-label use of rituximab, funding availability, risk tolerance, and convenience. This was evident in Norway, where differing preferences at two university hospitals in 2018 and 2019 created a natural experiment with two parallel cohorts: one treated with rituximab and the other with cladribine. This historical regional difference in MS treatment strategies, combined with a nationwide MS registry capturing data from over 85% of patients with MS, provided a unique opportunity to evaluate real-world long-term treatment effectiveness in two comparable cohorts.

To compare the effectiveness and safety of rituximab and cladribine, we emulated a target trial, a methodological approach to explicitly target the parameters from randomized trials when analyzing observational data that improve transparency and minimize the chance of self-inflicted biases in observational studies^6^. The primary outcome was MRI disease activity and the secondary outcomes included relapses, disability progression, and safety, with a median follow-up of 4.5 years.

## METHODS

### Design

We assessed treatment effectiveness in patients with RRMS using two population-based parallel cohorts, naturally created from two Norwegian university hospitals with different treatment strategies. Haukeland University Hospital (HUH) in Bergen predominantly treated patients with off-label rituximab, while Oslo University Hospital (OUH) in Oslo primarily used on-label cladribine. Since these two hospitals were exclusive healthcare providers for distinct geographical regions, treatment decisions for individual patients were largely determined by their residential address rather than personal factors. We used a target trial emulation framework to design the study, with the key protocol components outlined in Table 1.

**TABLE 1.**
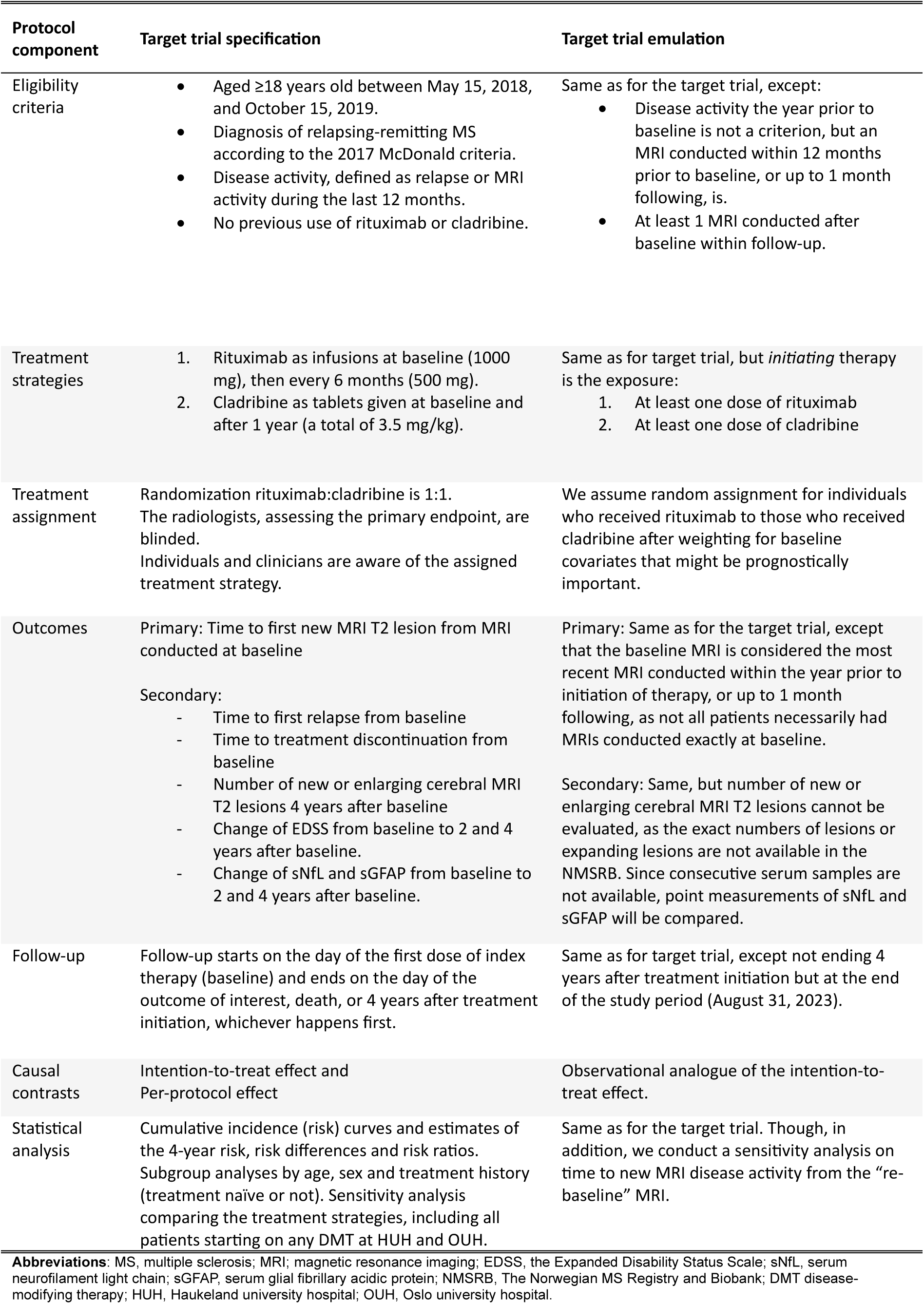
Specification and emulation of the target trial.

| Protocol component | Target trial specification | Target trial emulation |
| --- | --- | --- |
| Eligibility criteria | Aged $\geq 18$ years old between May 15, 2018, and October 15, 2019.<br>Diagnosis of relapsing-remitting MS according to the 2017 McDonald criteria.<br>Disease activity, defined as relapse or MRI activity during the last 12 months.<br>No previous use of rituximab or cladribine. | Same as for the target trial, except:<br>Disease activity the year prior to baseline is not a criterion, but an MRI conducted within 12 months prior to baseline, or up to 1 month following, is.<br>At least 1 MRI conducted after baseline within follow-up. |
| Treatment strategies | Rituximab as infusions at baseline (1000 mg), then every 6 months (500 mg).<br>Cladribine as tablets given at baseline and after 1 year (a total of 3.5 mg/kg). | Same as for target trial, but *initiating* therapy is the exposure:<br>At least one dose of rituximab<br>At least one dose of cladribine |
| Treatment assignment | Randomization rituximab:cladribine is 1:1.<br>The radiologists, assessing the primary endpoint, are blinded.<br>Individuals and clinicians are aware of the assigned treatment strategy. | We assume random assignment for individuals who received rituximab to those who received cladribine after weighting for baseline covariates that might be prognostically important. |
| Outcomes | Primary: Time to first new MRI T2 lesion from MRI conducted at baseline<br>Secondary:<br>Time to first relapse from baseline<br>Time to treatment discontinuation from baseline<br>Number of new or enlarging cerebral MRI T2 lesions 4 years after baseline<br>Change of EDSS from baseline to 2 and 4 years after baseline.<br>Change of sNfL and sGFAP from baseline to 2 and 4 years after baseline. | Primary: Same as for the target trial, except that the baseline MRI is considered the most recent MRI conducted within the year prior to initiation of therapy, or up to 1 month following, as not all patients necessarily had MRIs conducted exactly at baseline.<br>Secondary: Same, but number of new or enlarging cerebral MRI T2 lesions cannot be evaluated, as the exact numbers of lesions or expanding lesions are not available in the NMSRB. Since consecutive serum samples are not available, point measurements of sNfL and sGFAP will be compared. |
| Follow-up | Follow-up starts on the day of the first dose of index therapy (baseline) and ends on the day of the outcome of interest, death, or 4 years after treatment initiation, whichever happens first. | Same as for target trial, except not ending 4 years after treatment initiation but at the end of the study period (August 31, 2023). |
| Causal contrasts | Intention-to-treat effect and Per-protocol effect | Observational analogue of the intention-to-treat effect. |
| Statistical analysis | Cumulative incidence (risk) curves and estimates of the 4-year risk, risk differences and risk ratios.<br>Subgroup analyses by age, sex and treatment history (treatment naïve or not). Sensitivity analysis comparing the treatment strategies, including all patients starting on any DMT at HUH and OUH. | Same as for the target trial. Though, in addition, we conduct a sensitivity analysis on time to new MRI disease activity from the “re-baseline” MRI. |

**Abbreviations:** MS, multiple sclerosis; MRI; magnetic resonance imaging; EDSS, the Expanded Disability Status Scale; sNfL, serum neurofilament light chain; sGFAP, serum glial fibrillary acidic protein; NMSRB, The Norwegian MS Registry and Biobank; DMT disease-modifying therapy; HUH, Haukeland university hospital; OUH, Oslo university hospital.

### Study population

Patients included were identified from the Norwegian MS Registry and Biobank (NMSRB), with clinical and imaging data collected from routine patient care follow-ups. Data from the patient records were reviewed to ensure complete registration of covariates, exposures and outcomes in the registry prior to data extraction. Since adverse events were infrequently reported in the NMSRB, hospitalizations were separately registered from patients’ hospital records by experienced neurologists. In addition, serum samples from a nationwide COVID-19 vaccine response study^7,8^ were included and assessed for biomarker analyses.

Patients were eligible if they were ≥18 years old, had a diagnosis of RRMS, and had received at least one treatment with either cladribine at OUH or rituximab at HUH between May 15, 2018, and October 15, 2019. Patients were excluded if they had a progressive MS disease, were previously treated with either drug, or lacked reported data on baseline or consecutive MRIs. An individual’s baseline was defined as the date of initiation of treatment with cladribine or rituximab. The most recent MRI conducted within 12 months prior to baseline, or up to 1 month following, was considered the baseline MRI. The data cut-off date, also representing end of follow-up, was August 31, 2023. Due to harmonization efforts in Norway in 2015 and 2016, the MRI protocols for evaluating MS at the two centers were similar^9^.

### Exposures

All patients were treated (exposed) with either rituximab or cladribine at baseline, hereafter referred to as index therapy. The standard rituximab dosing at HUH was 1000 mg iv. at initiation, followed by 500 mg every 6 months^10^. The standard regimen of cladribine tablets was a cumulative dose of 3.5 mg/kg, over the first 2 years with cycles of 4 to 5 days twice each year^4^.

### Outcomes

The primary outcome was the time to new MRI disease activity, defined as new T2-lesions on brain or spinal cord MRI compared to baseline MRI. In the absence of competing events, cumulative incidence was estimated using a Kaplan-Meier estimator. Secondary outcomes were time to the first relapse, defined as an acute or subacute episode with symptoms and findings typical of MS, lasting at least 24 hours, with or without recovery, in the absence of infection or fever^11^; time to discontinuation of treatment or a third dose of cladribine; change in Expanded Disability Status Scale (EDSS) score; and the proportion of patients with no evidence of disease activity (NEDA-3), defined as no MRI disease activity, no relapse and no worsening in EDSS score. Additional secondary outcomes were serum levels of neurofilament light chain (NfL) and glial fibrillary acidic protein (GFAP) in the last available sample from each patient, measured with the Quanterix Simoa Neurology 2-Plex B Assay Kit; and hospitalizations related to possible adverse events (infections, malignancies and cardiac arrythmias) and deaths during index therapy. Multiple hospitalizations associated with the same type of adverse event in one patient were counted multiple times, except for malignancies, which were orderly counted as one event.

### Covariates

Baseline covariates included in the adjusted analyses were: age; sex; disease duration; number of previous disease modifying therapies (DMTs); total number of T2-lesions; EDSS score; relapses within 12 months prior to baseline; MRI lesion activity within 12 months prior to baseline; time between the most recent MRI conducted and baseline; and reasons for discontinuing the last DMT prior to baseline (Table 2).

**TABLE 2.** Baseline characteristics.

| Characteristic | No. (%) |  |
| --- | --- | --- |
|  | Cladribine<br>N = 126 | Rituximab<br>N = 159 |
| Age, mean (SD), y | 41 (11) | 42 (11) |
| Sex |  |  |
| Female | 95 (75) | 118 (74) |
| Male | 31 (25) | 41 (26) |
| Disease duration, median (IQR), y | 8 (11) | 6 (13) |
| No. of previous DMTs |  |  |
| 0 | 35 (28) | 73 (46) |
| 1 | 42 (33) | 31 (19) |
| 2 | 25 (20) | 31 (19) |
| 3 | 14 (11) | 14 (9) |
| 4 - 6 | 10 (8) | 10 (6) |
| MRI, T2 lesion load, n |  |  |
| 0 - 5 | 21 (17) | 20 (13) |
| 6 - 10 | 14 (11) | 46 (29) |
| >10 | 91 (72) | 93 (58) |
| Unknown | 0 (0) | 0 (0) |
| Disability, EDSS score<sup>a</sup> |  |  |
| Mild, 0 - 2 | 32 (25) | 40 (25) |
| Moderate, 2.5 - 5 | 19 (15) | 15 (9) |
| Severe, 5.5 and higher | 1 (1) | 5 (3) |
| Unknown | 74 (59) | 99 (62) |
| Relapse within 12 months before baseline | 46 (37) | 87 (55) |
| MRI disease activity within 12 months before baseline | 96 (76) | 110 (69) |
| Reason for discontinuing the last DMT before baseline |  |  |
| No DMTs prior to index treatment | 35 (28) | 73 (46) |
| Treatment failure | 30 (24) | 37 (23) |
| Side effects | 28 (22) | 29 (18) |
| Other reasons<sup>b</sup> | 32 (25) | 20 (13) |
| Unknown | 1 (1) | 0 (0) |
| Time between the most recent MRI and baseline, d | 57 (63) | 31 (58) |

<sup>a</sup> The last reported EDSS score within the year prior baseline.
<sup>b</sup> Patient's decision, family planning and other reasons.
**Abbreviations:** y, years; no., number; d, days; DMT, disease modifying therapy; MRI, magnetic resonance imaging; mo, months; EDSS, Expanded Disability Status Scale.

### Subgroup and sensitivity analyses

For the primary outcome, time to new MRI disease activity, we conducted prespecified subgroup analyses according to the treatment history (treatment-naïve or not), age (<40 or ≥40 years) and sex. As a prespecified sensitivity analysis, we compared the primary outcome between all patients at the two university hospitals who initiated any DMT within the specified baseline time interval. In this analysis, index therapy was defined as the first DMT started during the baseline interval. In addition, we included an analysis on the primary outcome from the re-baseline MRI. The re-baseline MRI was defined as the first MRI conducted after the baseline MRI, usually within 3-6 months after treatment initiation.

### Statistical Analysis

The occurrence of outcome events were described using plots of cumulative incidence (risk). In the absence of competing events, as no patients died, cumulative incidence was estimated using the Kaplan-Meier estimator. Absolute and relative risks differences between groups were estimated at 6, 2 years and 4 years. Adjustments for baseline covariates were performed using stabilized inverse probability of treatment (propensity score) weights, estimated using logistic regression^12^. Given the assumptions that the included baseline covariates sufficiently adjust for confounding, and that there is positivity (overlap) between the two treatment groups, group comparisons using the weighted data will correspond to the average treatment effect (ATE) of initiating rituximab versus initiating cladribine in the eligible patient population. Such effects of initiating treatments is sometimes referred to as an observational analogue of an intention-to-treat effect^12^. The treatment groups were compared on adjusted mean time free of new MR disease activity over 4 years (the weighted restricted mean survival time, RMST, and weighted log-rank test for difference)^13^. Percentile-based 95% confidence intervals (CI) for all estimates were calculated using non-parametric bootstrap with 1000 bootstrap samples. Baseline covariate balance was assessed using absolute standardized mean differences between the groups, with a difference of up to 0.1 considered acceptable. For the sensitivity analysis on MRI lesion activity after the re-baseline MRI, we estimated the hazard ratio using the weighted Cox proportional hazard model.

To estimate change in EDSS score per year, the most recent EDSS score registered up to 12 months prior to baseline was compared with the last EDSS score registered within the observational time. One or both EDSS scores were missing for 184 (64%) patients, and representativeness was analyzed by comparing baseline characteristics. EDSS change was compared using the Wilcoxon rank sum test. Proportions with NEDA-3 status were compared using the Chi-square test. NfL and GFAP levels were first log-transformed, then compared between the treatment groups, using the Student’s two-sample *t*- test and a linear-regression analysis, adjusting for sex and age. The confidence intervals were calculated at the 2-sided 95% level.

The statistical analyses were conducted using R, version 4.1.2 (R Foundation for Statistical Computing), with the packages cobalt, WeightIt and survival. Figures were created in R using ggplot2.

### Standard Protocol Approvals, Registrations, and Patient Consents

The Norwegian Regional Committee for Medical Research Ethics approved the study (REK no 629865). All patients had provided written informed consent to participate in the NMSRB for research purposes. The Strengthening the Reporting of Observational Studies in Epidemiology (STROBE) reporting guidelines for cohort studies were followed in the reporting of this study.

### Data availability

Data can be obtained upon request. Inquiries should be directed to the Norwegian MS Registry and Biobank, which follows a protocol for evaluating data access requests. Due to privacy regulations, the data cannot be made publicly available in a repository.

## RESULTS

### Study population and follow-up

We included 285 patients with RRMS; 159 patients were treated with rituximab and 126 were treated with cladribine (Figure 1). Baseline characteristics are presented in Table 2. After inverse probability weighting, covariates were balanced between the treatment groups, except in patients with severe disability and those with “other reasons” for discontinuing the previous DMT (eFigure 1). The median follow-up period was 4.5 years (interquartile range, IQR: 4.0 to 5.0) and most patients were censored at the end of the study observation period. A median of 5 MRIs (IQR: 4 to 6) were conducted between baseline and the end of the study observation period for both treatment groups.

**FIGURE 1.**
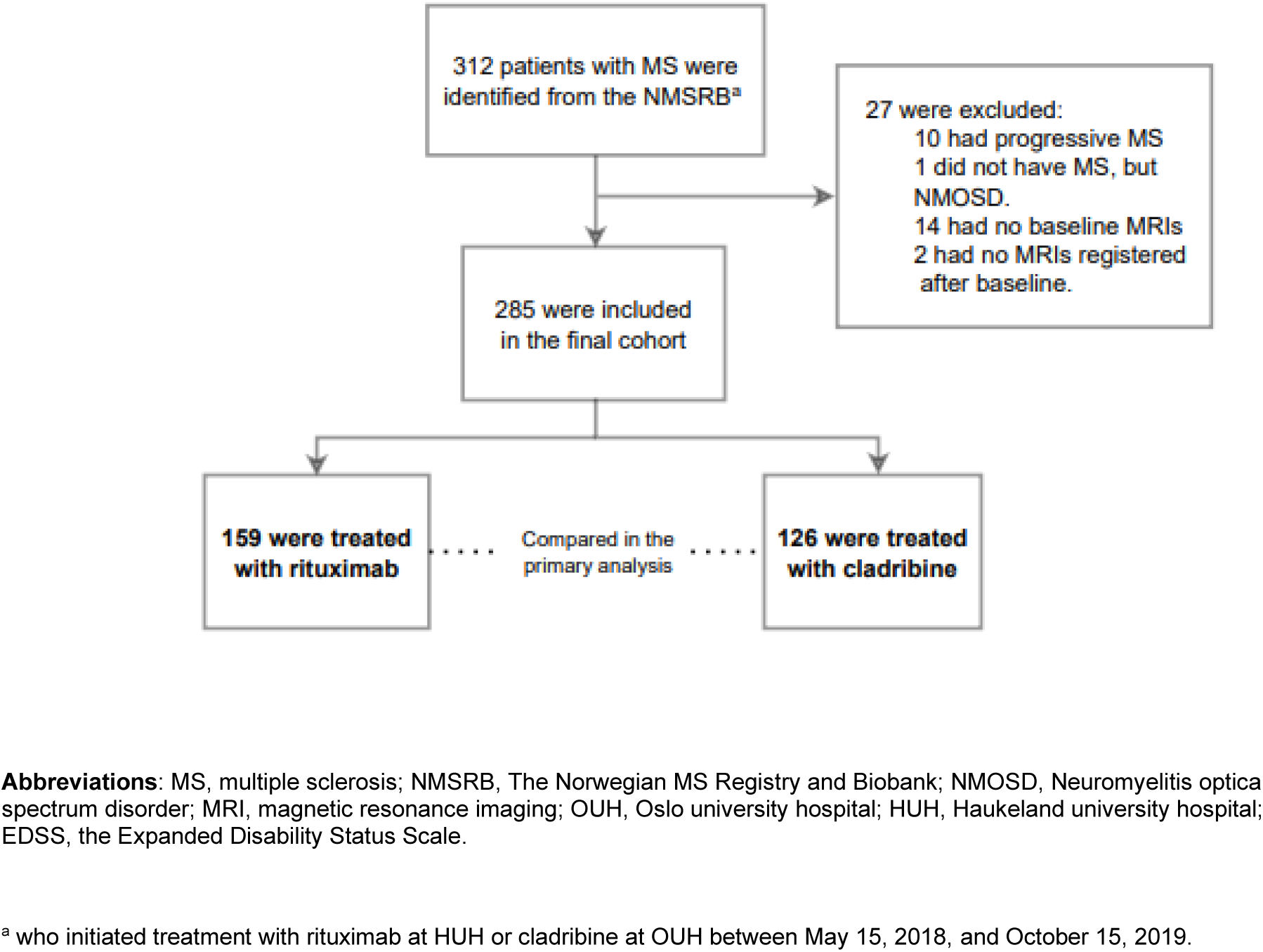
Flowchart of study inclusion.

### Primary outcome – new MRI disease activity

The cumulative incidence curve for new MRI disease activity is shown in Figure 2. Patients treated with rituximab had a lower risk of new MRI disease activity during follow-up compared to patients treated with cladribine (weighted log-rank test, *p* < 0.0001). The 4-year risk of new MRI disease activity was 18% (95% CI 11 to 23) for patients treated with rituximab and 57% (95% CI 48 to 65) for patients treated with cladribine. The risk difference (RD) at 6 months, 2 years and 4 years after treatment initiation were 22.6 percentage-points (95% CI 15 to 24), 32.7 percentage-points (95% CI 23 to 43) and 38.1 percentage-points (95% CI 29 to 51). The risk ratio (RR) at 6 months, 2 years and 4 years after treatment initiation were 0.34 (95% CI 0.19 to 0.51), 0.32 (95% CI 0.20 to 0.46) and 0.31 (95% CI 0.20 to 0.43) for patients treated with rituximab compared to cladribine (eTable 1 and 2 and Table 3). During the 4.5 years of follow-up, the patients treated with rituximab were free of new MRI disease activity for a mean of 16.2 months (95% CI 9.9 to 23.2) longer than those treated with cladribine.

**FIGURE 2.**
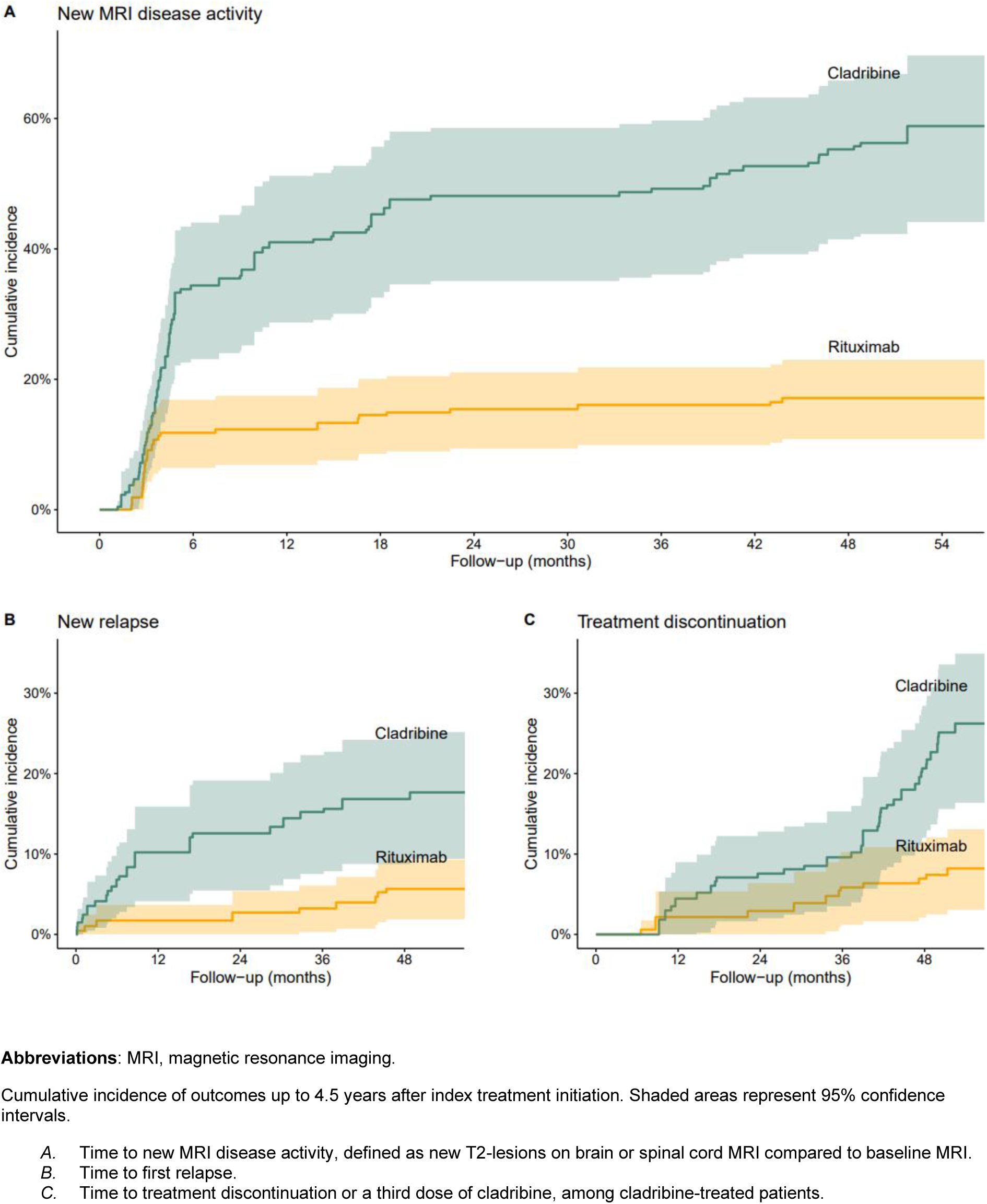
Cumulative incidence of outcomes.

**TABLE 3.** Comparative effectiveness 4 years after initiation of rituximab (n= 159) and cladribine (n = 126) in patients with multiple sclerosis.

| Outcome | Number of events |  | 4-year risk (95% CI) |  | Risk difference<sup>c</sup><br>(95% CI) | Risk ratio<sup>c</sup><br>(95% CI) |
| --- | --- | --- | --- | --- | --- | --- |
|  | Rituximab | Cladribine | Rituximab | Cladribine |  |  |
|  |  |  |  |  | *percentage-points* |  |
| New MRI activity<sup>a</sup> | 28 | 72 | 18%<br>(11 to 23) | 57%<br>(48 to 65) | 38.1<br>(29.0 to 51.0) | 0.31<br>(0.20 to 0.43) |
| New relapse | 9 | 21 | 5.7%<br>(2.0 to 9.2) | 17%<br>(9.9 to 23) | 11.2<br>(3.1 to 18.1) | 0.34<br>(0.13 to 0.67) |
| Treatment discontinuation<sup>b</sup> | 9 | 28 | 5.7%<br>(2.0 to 9.2) | 22%<br>(15 to 29) | 13.7<br>(9.0 to 25.0) | 0.34<br>(0.10 to 0.48) |

<sup>a</sup> New MRI disease activity, defined as new T2-lesions on brain or spinal cord MRI compared to baseline MRI.
<sup>b</sup> Treatment discontinuation or third dose of cladribine, among cladribine-treated patients.
<sup>c</sup> Adjusted for age; sex; disease duration; number of previous disease modifying therapies (DMTs); number of T2-lesions on MRI; EDSS score; relapses within 12 months prior to baseline; MRI lesion activity within 12 months prior to baseline; time from baseline MRI to baseline; and reasons for discontinuing the last DMT prior to the index therapy.
**Abbreviations:** CI, confidence interval; MRI, magnetic resonance imaging.

### Secondary outcomes

The risk of relapses 4 years after treatment initiation was lower for patients treated with rituximab (5.7% [95% CI 2.0 to 9.2]) than for patients treated with cladribine (17% [95% CI 9.9 to 23], *p* = 0.0012), with an RD of 11.2 percentage-points (95% CI 3 to 18) and an RR of 0.34 (95% CI 0.13 to 0.67; Table 3). After 4 years, fewer rituximab-treated patients had discontinued their therapy (5.7% [95% CI 2.0 to 9.2]) compared to those treated with cladribine (22% [95% CI 15 to 29], *p* < 0.0001), with an RD of 13.7 percentage-points (95% CI 9 to 25) and an RR of 0.34 (95% CI 0.10 to 0.48). The main reasons for discontinuing index therapy or receiving a third dose of cladribine were side effects for rituximab and disease activity for cladribine (eTable 3). Nine cladribine-treated patients received a third dose, while 26 switched another DMT.

EDSS scores were available at baseline and during follow-up for 101 patients (35%). Baseline covariates were similar between those with and without EDSS scores available, except that there were fewer treatment-naïve patients with EDSS scores. The median EDSS score change was 0.0 per year in the rituximab-treated patients and 0.1 per year in the cladribine-treated patients (*p* < 0.001; eFigure 2). The proportion with NEDA-3 status was higher in rituximab-treated patients (58%) compared to cladribine-treated patients (14%, *p* < 0.0001; eFigure 2).

NfL and GFAP were available for analysis in 133 patients (47%) from serum samples collected 1.7 to 4.2 years after baseline. NfL levels did not differ between the treatment groups (5.6 pg/mL in the rituximab-treated patients vs. 6.9 pg/mL in the cladribine-treated patients, *p* = 0.10). GFAP levels were lower in the rituximab-treated patients (62.6 pg/mL) compared to the cladribine-treated patients (87.8 pg/mL, *p* = 0.013; eFigure 3), and the difference persisted after adjusting for age and sex (*p* = 0.02).

Information about adverse events was available for 284 of 285 patients (99.6%). The incidence of hospitalizations related to possible adverse events during rituximab therapy was 6.0 per 100 person-years versus 4.1 per 100 person-years during cladribine therapy. The most frequent reason for hospitalization was COVID-19 (including post-acute COVID-19 syndrome) in the rituximab-treated patients with the highest incidence in 2021 to 2022, while in the cladribine-treated patients, it was other respiratory diseases (Table 4 and eTable 4). No patients died during the follow-up period.

**TABLE 4.** Hospitalizations related to possible adverse events.

|  | Therapy |  |
| --- | --- | --- |
|  | Rituximab<br>N = 158<br>697 PY | Cladribine<br>N = 126<br>516 PY |
| Hospitalizations due to any possible adverse drug event, n | 42 | 21 |
| Patients with $\geq 1$ hospitalization due to any possible adverse drug event, n | 26 | 12 |
| Hospitalizations per 100 person-years | 6.0 | 4.1 |
| **Adverse events per ICD-10 categories, n** |  |  |
| Infectious and parasitic diseases | 3 | 3 |
| COVID-19 and post-acute COVID-19 syndrome | 16 | 2 |
| Diseases of the circulatory system | 4 | 2 |
| Diseases of the digestive system | 0 | 2 |
| Diseases of the genitourinary system | 3 | 4 |
| Diseases of the musculoskeletal system and connective tissue | 0 | 1 |
| Diseases of the nervous system | 0 | 1 |
| Diseases of the respiratory system | 13 | 5 |
| Injury, poisoning and certain other consequences of external causes | 1 | 0 |
| Neoplasms | 1 | 1 |
| Symptoms, signs and abnormal clinical and laboratory findings, not elsewhere classified | 1 | 0 |

Multiple hospitalizations for the same type of adverse event in one patient were counted multiple times, except for malignancies, which were orderly counted as one event.
**Abbreviations:** PY, person-years; ICD, International Classification of Diseases.

### Subgroup and sensitivity analysis

The cumulative incidence curves were similar across subgroups defined by treatment history, age and sex (eFigure 4). The re-baseline MRIs were conducted a median of 3.1 months after the initiation of rituximab and 3.6 months after the initiation of cladribine. Analyses including data from the re-baseline MRI showed that patients who were treated with rituximab had a lower risk of new MRI disease activity, compared to those who were treated with cladribine, with a hazard ratio of 0.15 (95% CI 0.07 to 0.30, *p* < 0.0001; eFigure 5).

When comparing new MRI disease activity between the two hospitals with different treatment strategies, including all patients regardless of type of initial DMT, we included 436 patients: 193 at HUH and 243 patients at OUH. At HUH, 80% of the patients started on rituximab and 1% started on cladribine, while at OUH, 49% started cladribine and 11% started rituximab (eTable 5). Patients treated at HUH had lower risk of new MRI disease activity, compared to patients treated at OUH (*p* < 0.0001, eFigure 6).

## DISCUSSION

In this study, we found that rituximab had superior effectiveness compared to cladribine in reducing new MRI disease activity, relapses, treatment discontinuation, and disability worsening over a median follow-up of 4.5 years. The incidence of hospitalizations potentially related to adverse events was similar between the two treatment groups.

Our results on new MRI disease activity, a sensitive and objective marker of disease activity routinely used in clinical practice^11,14^, were consistent with those reported from other studies. In the RIFUND- MS trial, which randomized treatment-naïve patients to rituximab or dimethyl fumarate, 21% of the rituximab-treated patients had new lesions on MRI after two years^3^, the same proportion as for our subgroup of treatment-naïve patients who received rituximab. In the placebo-controlled CLARITY and the CLARITY Extension trial^15, 16^, 65.6% of the patients who received two courses of cladribine had active T2 lesions in the following two years^16^. In our cohort, 57% (95% CI 48 to 65) had new lesions 4 years after cladribine initiation. An ongoing randomized trial (NOR-MS, NCT04121403) is comparing rituximab and cladribine over a two-year of follow-up period, with new MRI T2-lesions as the primary outcome. Results are expected to complement our findings.

The marked difference in the risk of MRI disease activity that emerged within the first 6 months between rituximab and cladribine suggests a delayed treatment effect of cladribine. Rapid treatment effect is particularly crucial when there is a high risk of continued or rebound disease activity, as described after discontinuation of fingolimod and natalizumab^17,18^. We have previously reported a higher risk of rebound among patients who switched from fingolimod to cladribine (21%, 7/33), compared to none of the patients who switched to rituximab (0/40)^19^. However, the difference in effectiveness in our study was not limited to the first months after treatment initiation or to prior DMT, as the difference persisted in both the subgroup of treatment-naïve patients and when analyzing time to MRI disease activity from the re-baseline MRI.

The 4-year risk of relapse was lower for rituximab compared to cladribine in our study. This is consistent with a recent registry-based publication from the MSBase, which found a lower relapse rate among patients treated with ocrelizumab compared to those treated with cladribine (0.05 vs 0.09, p = 0.008)^20^. However, we observed a lower cumulative incidence of relapses for both rituximab and cladribine, compared to the previous RCTs^3–5^. This finding may be due to differences in patient composition, since the original rituximab and cladribine studies were performed a decade before the present study^4,5^, or to a more stringent use of the relapse definitions and differences in reporting of relapses, as suggested in a report from the Swedish MS registry^21^.

The changes in EDSS during follow-up in our study suggest no change in disability score in patients treated with rituximab compared to an overall slight worsening among those treated with cladribine, consistent with earlier findings^20^. Serum NfL, a biomarker of axonal damage and acute inflammation^22^, did not differ significantly between the treatment groups in our study. On the other hand, serum GFAP, an intermediate filament in astrocytes and a proposed biomarker for disease progression independent of relapse activity^22,23^, was found at lower levels in patients who were treated with rituximab compared to those who were treated with cladribine. While this finding correlates with the observed EDSS progression, it should be interpreted cautiously, as no longitudinal samples were available, and the role of serum GFAP in MS is not yet fully established.^23^

The observation period of our study included the years of the COVID-19 pandemic. During this time, infections were particularly concerning for patients treated with anti-CD20 antibodies, as they had an attenuated humoral vaccine response^7^. Although studies have reported a high relative risk of hospitalization and severe COVID-19 infection associated with anti-CD20 treatment compared to other DMTs^24, 25,26^, the absolute risk of severe COVID-19 infections in Norway has been low^27^. In our study, COVID-19 was the most frequent adverse event related to hospitalizations among the patients treated with rituximab. This may partly be related to an increased awareness and early healthcare response for these patients, where patients on rituximab were eligible for early antiviral treatment in case of acquired COVID-19 disease. The number of hospitalizations potentially related to adverse events other than COVID-19 was similar for patients treated with rituximab and cladribine in our study. This aligns with other reports that has indicated the same risk of infections for rituximab as other high-efficacy therapies^28^ and in the RCTs, where risks of serious infections were similar between the anti-CD20 treatment arms and the comparators (placebo, interferon-beta 1a, teriflunomide and dimethyl fumarate)^3,5,29–31^. However, a Swedish observational study reported a higher risk of serious infections in patients with MS treated with rituximab compared to those treated with natalizumab, fingolimod, interferon beta, and glatiramer acetate^32^. Future studies may shed light on the relative risk of infections with different DMTs.

### Strengths and limitations

Our study population emerged from a near-randomly occurring difference in treatment preferences between two hospitals, where treatment was primarily determined by patients’ residential addresses. While it is impossible to distinguishing treatment allocation from local assessment practices, Norway’s homogeneous population, standardized healthcare system, and national clinical guidelines for MS management likely minimize this issue. Additionally, the high-coverage Norwegian MS- registry provided a unique opportunity to assess real-world long-term effectiveness, including a broader age range and without excluding patients with comorbidities, knowledge that is usually not captured in RCTs. To further minimize common biases in observational studies, we applied the target trial framework, which allowed us to structure the study in a way that facilitates causal inference.

The main limitation of this study is the lack of randomization. We cannot account for all potential confounders, such as smoking status, body mass index, comorbidity, ethnic background or other unmeasured confounders. Nonetheless, the thorough sub-analyses and similarity of our estimates to those reported from previous RCTs, suggest that our findings are reliable. The scarcity of EDSS data patient records represents another limitation, and these results should be interpreted cautiously. However, they align with the MRI and relapse activity findings. Another limitation is that serum NfL and GFAP were only measured at one time point per patient, with samples collected randomly throughout the treatment course, preventing evaluation on changes from treatment start and during therapy. Finally, the time between the baseline MRI scan and the initiation of index therapy varied among the patients, and therefore, new lesions reported at the re-baseline MRI could have appeared before therapy initiation. However, the additional sensitivity analysis of the primary outcome, analyzing new MRI disease activity after the re-baseline MRI examination, confirmed the consistent pattern of difference in treatment efficacy between the groups.

In summary, rituximab showed superior effectiveness over cladribine on all measures of efficacy and in all subgroups in this observational population-based comparative study.

## Supplementary Appendix

Supplementary data to BE. Rød, EA. Høgestøl, Ø. Torkildsen, K. Bjørnevik, JM. Gran, MH. Øverås, M. König, EG. Celius, K-M Myhr, S. Wergeland, GO. Nygaard **Comparative Effectiveness of Rituximab and Cladribine in Relapsing-Remitting Multiple Sclerosis: A Target Trial Emulation**.

## FIGURES

**eFigure 1.**
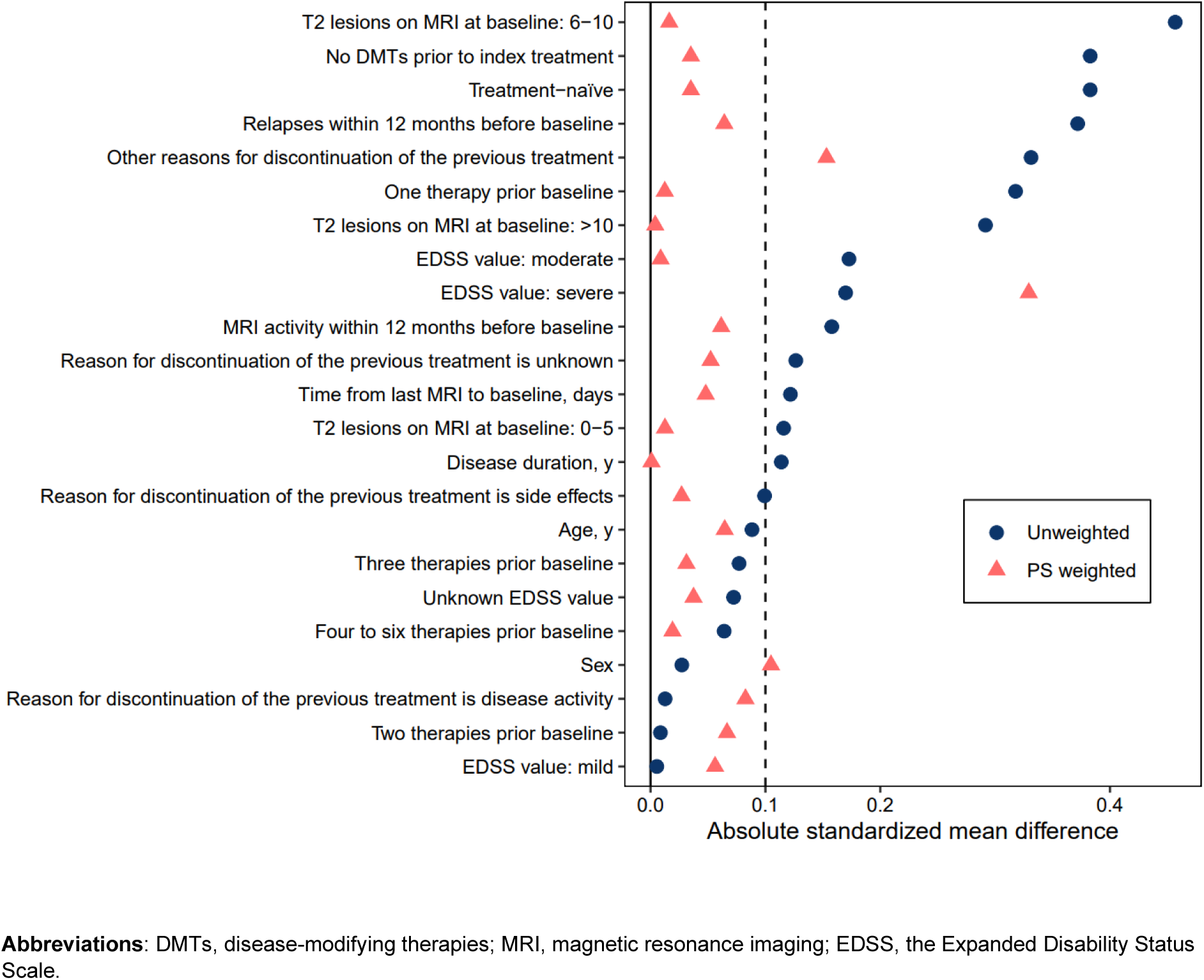
Absolute standardized mean differences before and after applying inverse probability weighting.

**eFigure 2.**
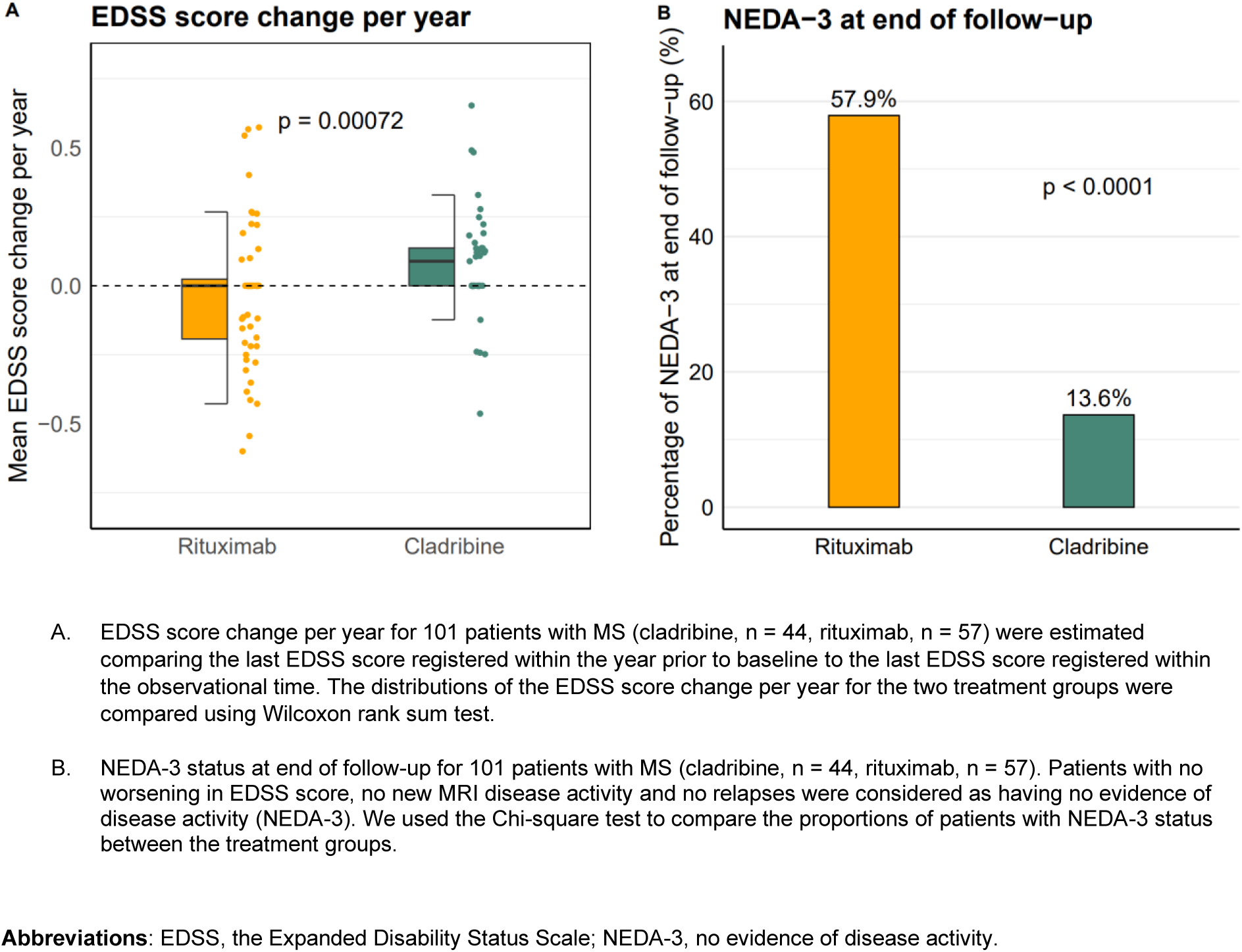
EDSS score change per year and proportions with NEDA-3 status at end of follow-up.

**eFigure 3.**
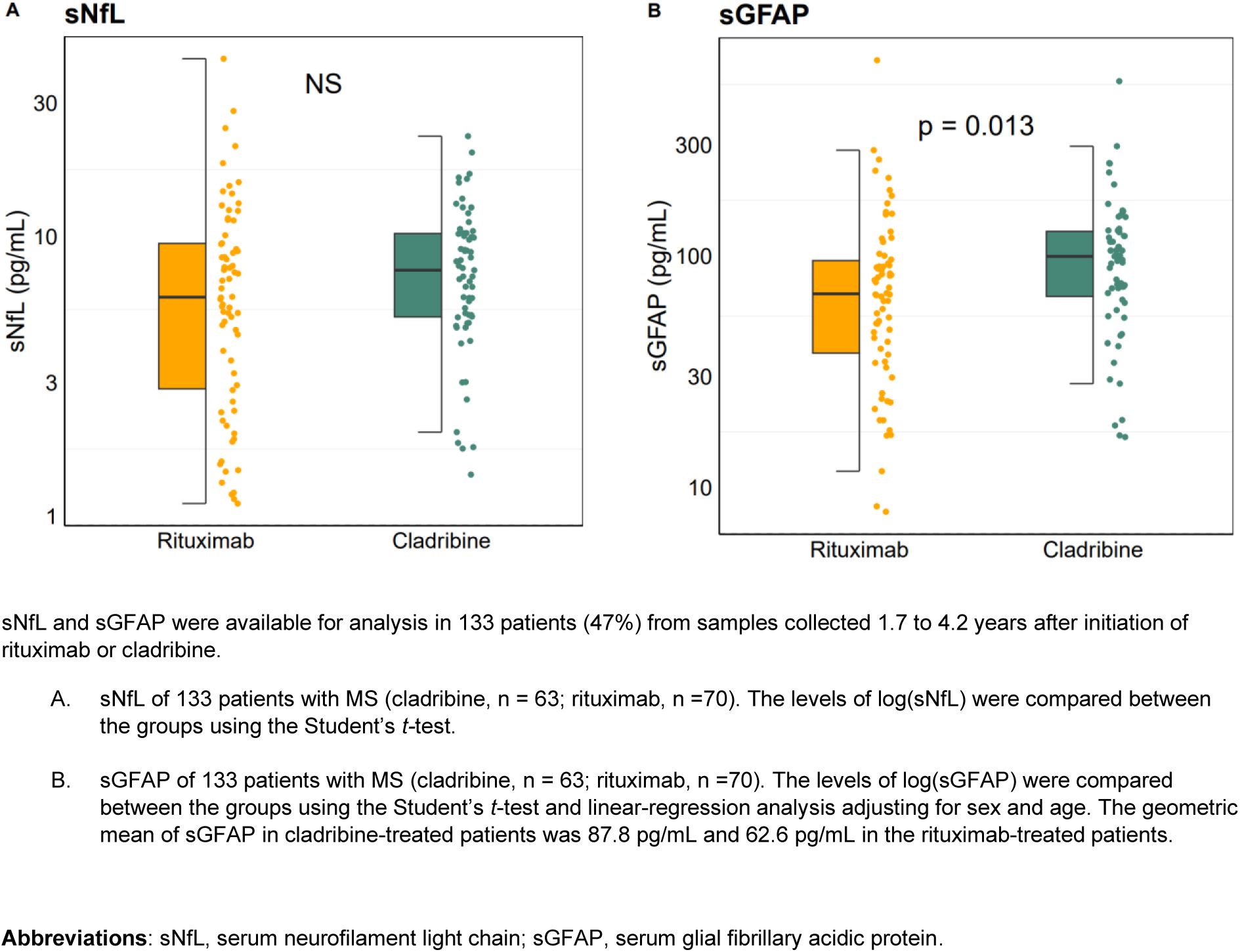
sNfL and sGFAP

**eFigure 4.**
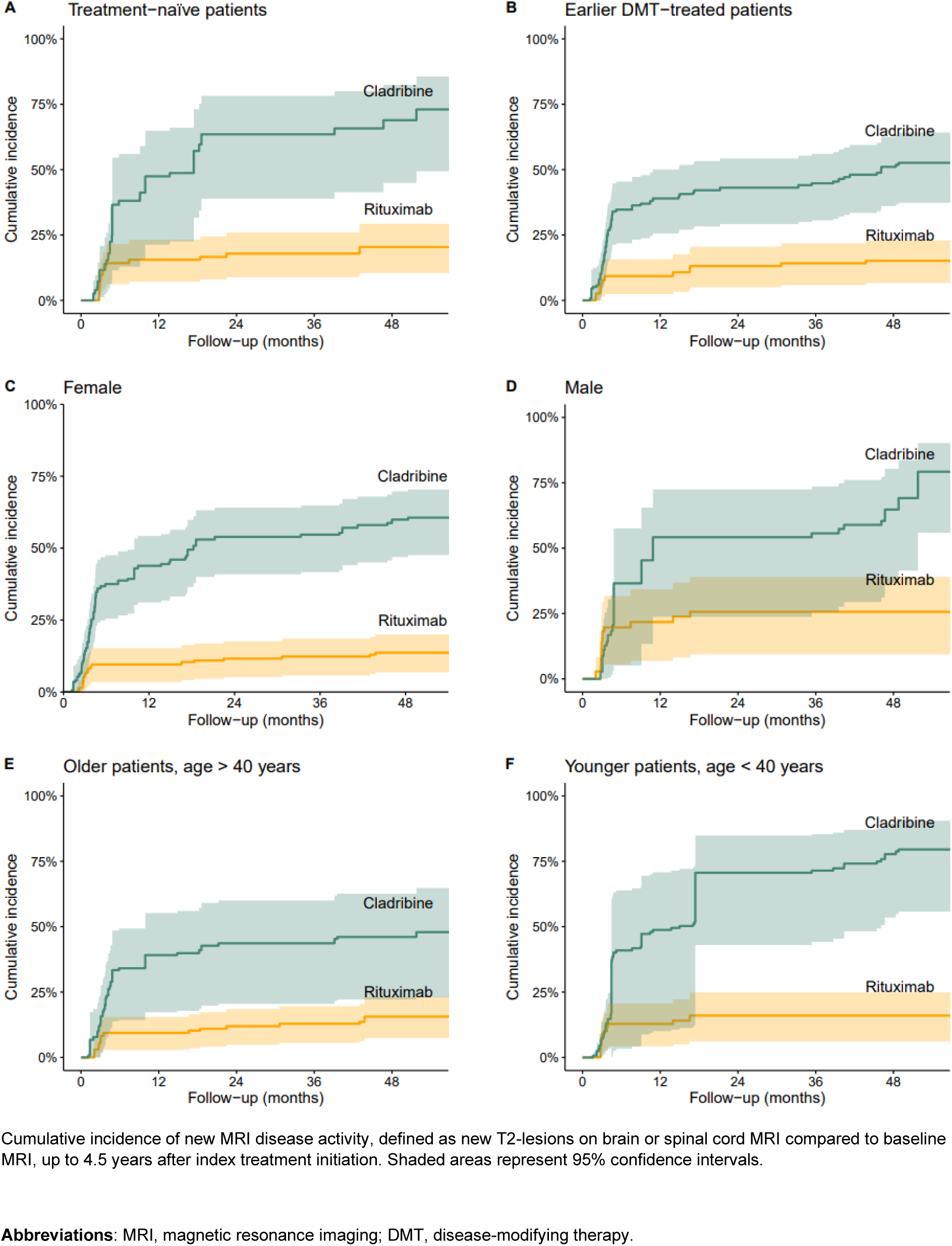
Cumulative incidence of new MRI disease activity in subgroups.

**eFigure 5.**
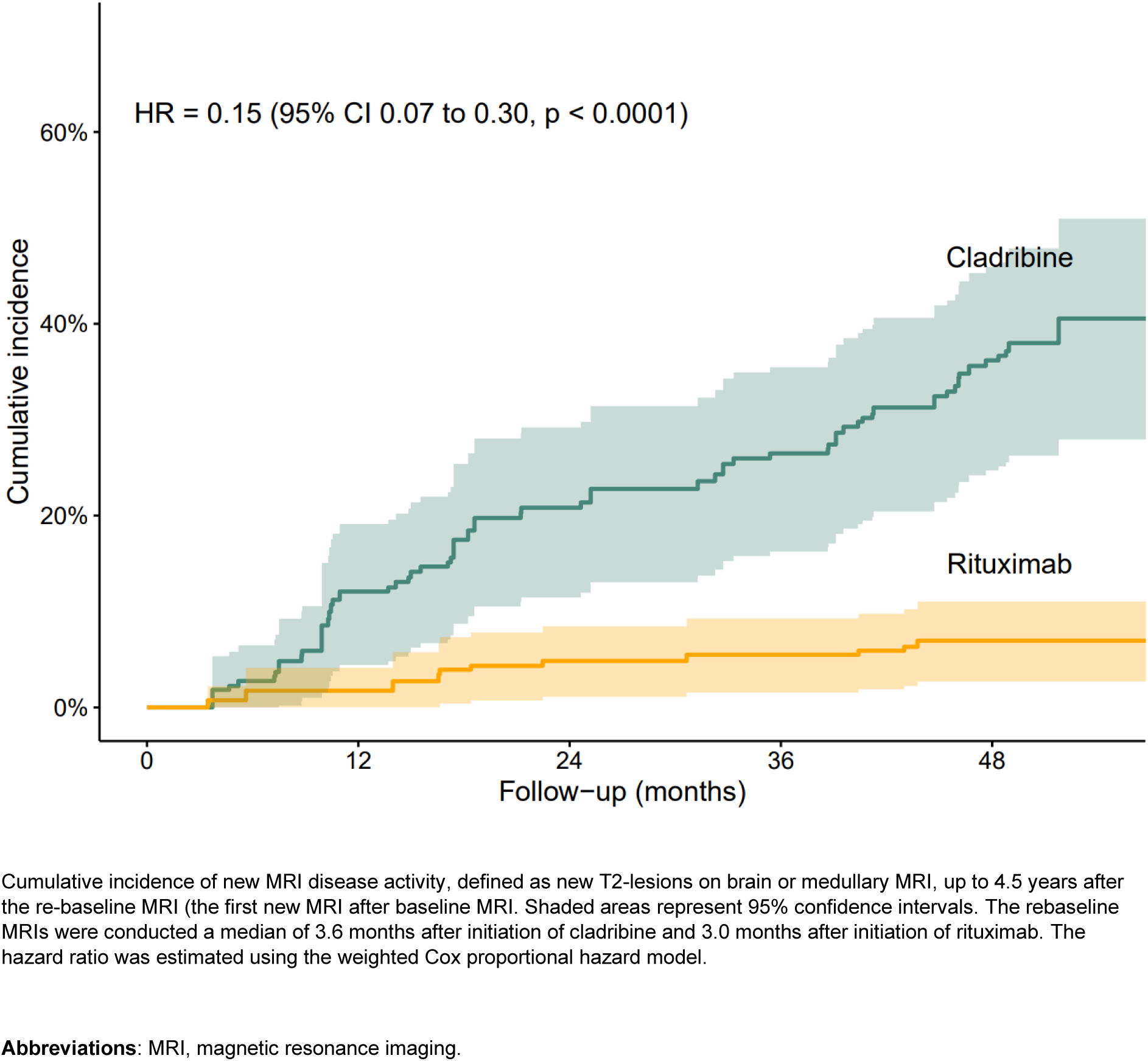
Cumulative incidence of new MRI disease activity from re-baseline

**eFigure 6.**
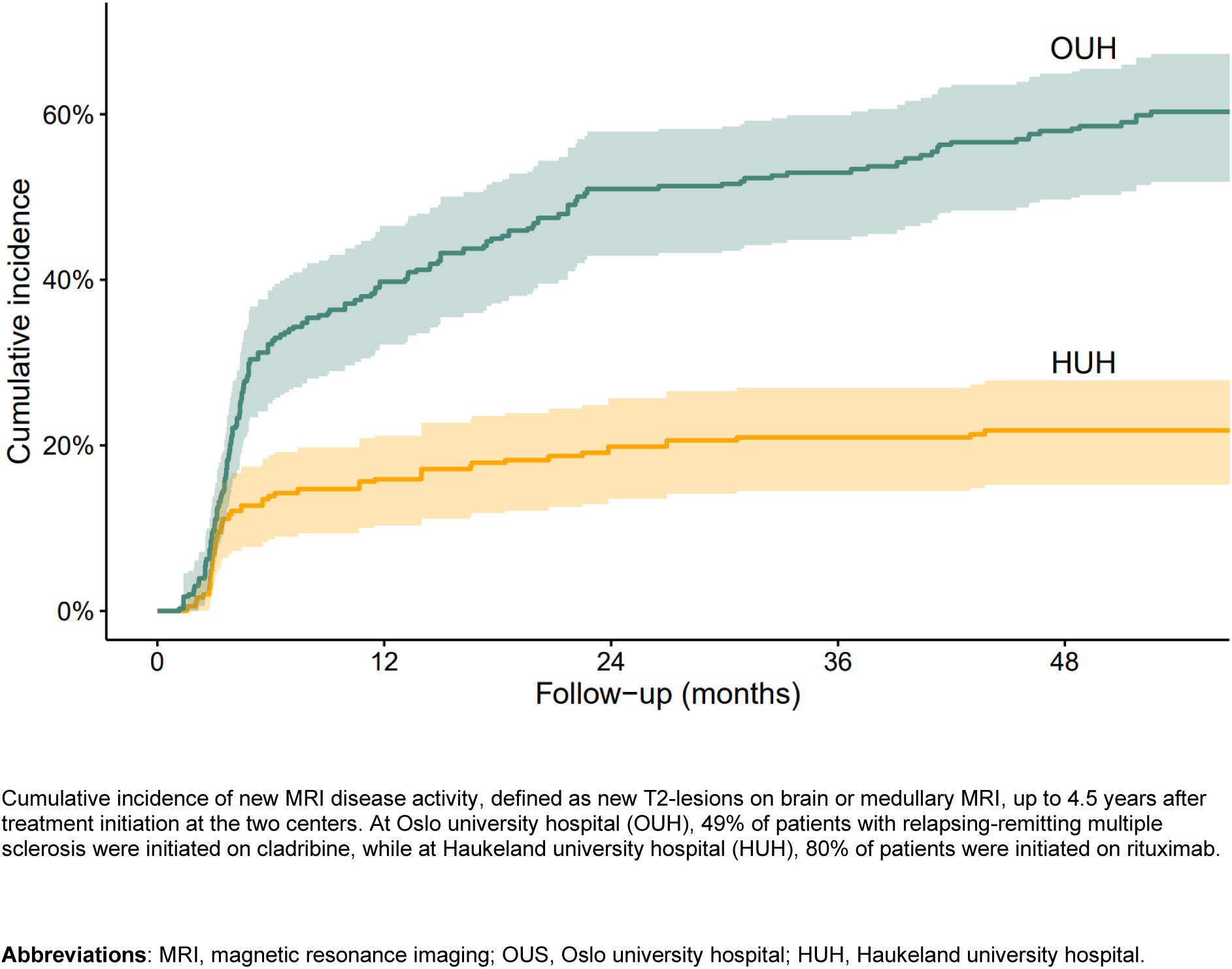
Cumulative incidence of new MRI disease activity by treatment strategy cohorts

## TABLES

**eTable 1.** Comparative effectiveness 6 months after initiation of rituximab (n= 159) and cladribine (n = 126) in patients with multiple sclerosis.

| Outcome | Number of events |  | 6-month risk (95% CI) |  | Risk difference<sup>c</sup><br>(95% CI) | Risk ratio<sup>c</sup><br>(95% CI) |
| --- | --- | --- | --- | --- | --- | --- |
|  | Rituximab | Cladribine | Rituximab | Cladribine |  |  |
|  |  |  |  |  | *percentage-points* |  |
| New MRI activity<sup>a</sup> | 19 | 46 | 12%<br>(6.8 to 17) | 37%<br>(28 to 44) | 22.6<br>(15.0 to 34.0) | 0.34<br>(0.19 to 0.51) |
| New relapse | 3 | 11 | 1.9%<br>(0 to 4.0) | 8.7%<br>(3.7 to 14) | 5.1<br>(2.0 to 12.4) | 0.25<br>(0.00 to 0.67) |
| Treatment discontinuation<sup>b</sup> | 0 | 0 | 0%<br>(0 to 0) | 0%<br>(0 to 0) | 0<br>(0 to 0) | NaN |

<sup>a</sup> New MRI disease activity, defined as new T2-lesions on brain or spinal cord MRI compared to baseline MRI.
<sup>b</sup> Treatment discontinuation or third dose of cladribine, among cladribine-treated patients.
<sup>c</sup> Adjusted for age; sex; disease duration; number of previous disease-modifying therapies (DMTs); number of T2-lesions on MRI; EDSS score; relapses within 12 months prior to baseline; MRI lesion activity within 12 months prior to baseline; time from baseline MRI to baseline; and reasons for discontinuing the last DMT prior to the index therapy.
**Abbreviations:** CI, confidence interval; MRI, magnetic resonance imaging.

**eTable 2.** Comparative effectiveness 2 years after initiation of rituximab (n= 159) and cladribine (n = 126) in patients with multiple sclerosis.

| Outcome | Number of events |  | 2-year risk (95% CI) |  | Risk difference<sup>c</sup><br>(95% CI) | Risk ratio<sup>c</sup><br>(95% CI) |
| --- | --- | --- | --- | --- | --- | --- |
|  | Rituximab | Cladribine | Rituximab | Cladribine |  |  |
|  |  |  |  |  | *percentage-points* |  |
| New MRI activity<sup>a</sup> | 25 | 61 | 16%<br>(9.9 to 21) | 48%<br>(39 to 56) | 32.7<br>(22.6 to 43.1) | 0.32<br>(0.20 to 0.46) |
| New relapse | 4 | 16 | 2.5%<br>(<0.1 to 4.9) | 13%<br>(6.7 to 18) | 9.9<br>(4.3 to 16.4) | 0.22<br>(0.04 to 0.47) |
| Treatment discontinuation<sup>b</sup> | 3 | 9 | 1.9%<br>(0 to 4.0) | 7.1%<br>(2.5 to 12) | 4.6<br>(0.4 to 10.6) | 0.39<br>(0.00 to 0.93) |

<sup>a</sup> New MRI disease activity, defined as new T2-lesions on brain or spinal cord MRI compared to baseline MRI.
<sup>b</sup> Treatment discontinuation or third dose of cladribine, among cladribine-treated patients.
<sup>c</sup> Adjusted for age; sex; disease duration; number of previous disease-modifying therapies (DMTs); number of T2-lesions on MRI; EDSS score; relapses within 12 months prior to baseline; MRI lesion activity within 12 months prior to baseline; time from baseline MRI to baseline; and reasons for discontinuing the last DMT prior to the index therapy.
**Abbreviations:** CI, confidence interval; MRI, magnetic resonance imaging.

**eTable 3.** Reason for discontinuation or third dose of cladribine.

| Reason for discontinuation | No. (%) |  |
| --- | --- | --- |
|  | Rituximab<br>N = 11 | Cladribine<br>N = 35 |
| Disease activity | 2 (18) | 24 (69) |
| Adverse events | 6 (55) | 1 (3) |
| Other reasons<sup>a</sup> | 3 (27) | 2 (6) |
| Unknown | 0 (0) | 8 (23) |

<sup>a</sup>include family planning, patient's decision and other reasons.

**eTable 4.** Timing of adverse events after treatment initiation.

| **Hospitalizations related to adverse events, per 100 patient-years**<br>(Number of hospitalizations / patient years of observation on DMT monotherapy) |  |  |
| --- | --- | --- |
|  | **Rituximab**<br>**N = 158** | **Cladribine**<br>**N = 126** |
| Year 1 | **6.4**<br>(10/157.3) | **2.4**<br>(3/125.5) |
| Year 2 | **2.6**<br>(4/155.8) | **5.0**<br>(6/119.5) |
| Year 3 | **1.3**<br>(2/154.1) | **5.2**<br>(6/116.3) |
| Year 4<sup>a</sup> | **11.3**<br>(17/149.8) | **4.6**<br>(5/109.7) |
| Year 5<sup>b</sup> | **11.7**<br>(9/76.7) | **2.3**<br>(1/44.4) |
| Year 6 | **0**<br>(0/2.8) | **0**<br>(0/0.4) |
| **Total** | **6.0**<br>(42/696.5) | **4.1**<br>(21/515.4) |

<sup>a</sup> Corresponding to 2021-2022. <sup>b</sup> Corresponding to 2022-2023.
**Abbreviations:** DMT, disease-modifying therapy.

**eTable 5.**
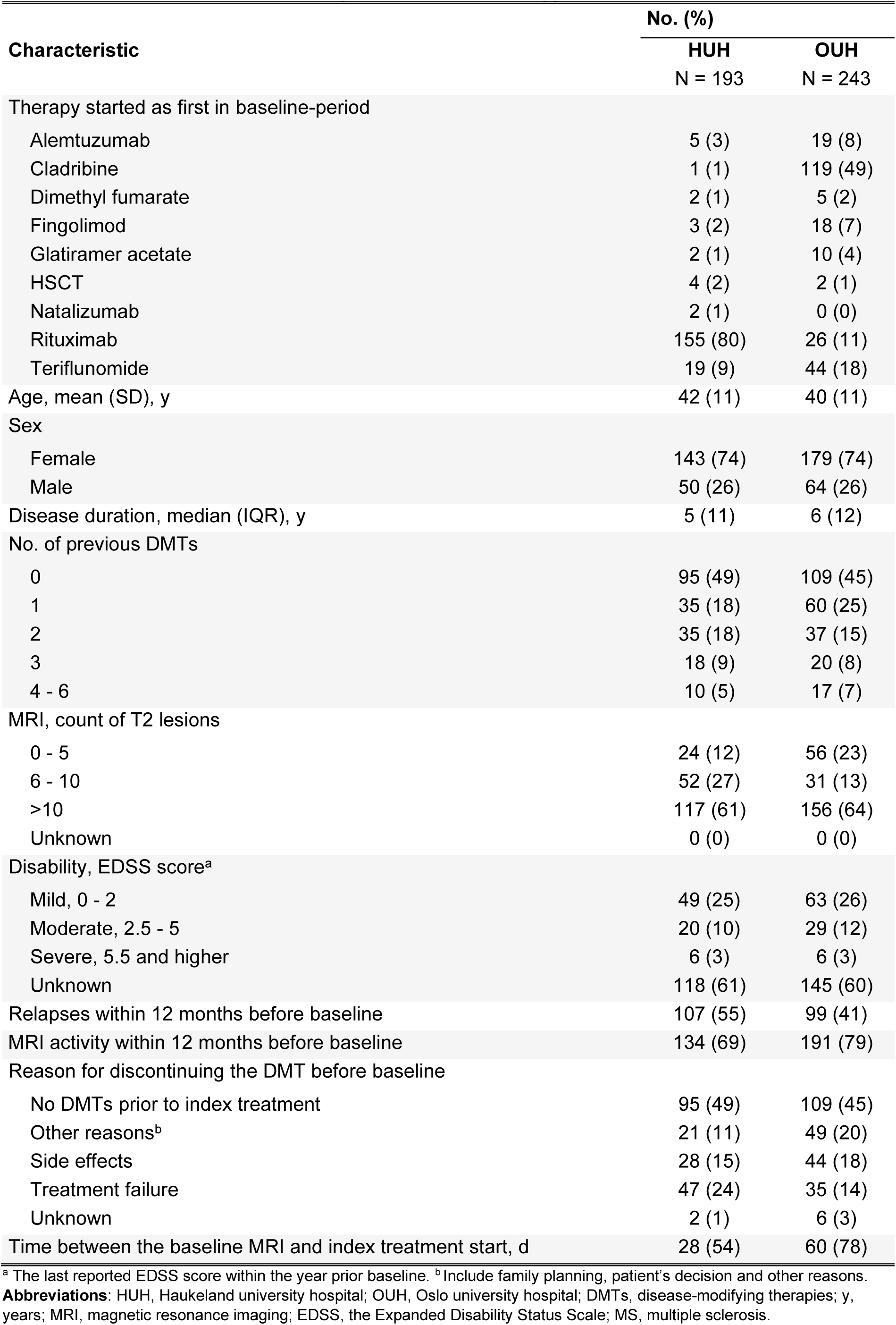
Baseline variables by treatment strategy cohort.

## References

1. Freeman L, Longbrake EE, Coyle PK, Hendin B, Vollmer T. High-Efficacy Therapies for Treatment-Naïve Individuals with Relapsing-Remitting Multiple Sclerosis. CNS Drugs. Dec 2022;36(12):1285–1299. doi:10.1007/s40263-022-00965-7

2. Henderson M, Horton DB, Bhise V, Pal G, Bushnell G, Dave CV. Initiation Patterns of Disease-Modifying Therapies for Multiple Sclerosis Among US Adults and Children, 2001 Through 2020. JAMA Neurol. Aug 1 2023;80(8):860–867. doi:10.1001/jamaneurol.2023.2125

3. Svenningsson A, Frisell T, Burman J, et al. Safety and efficacy of rituximab versus dimethyl fumarate in patients with relapsing-remitting multiple sclerosis or clinically isolated syndrome in Sweden: a rater-blinded, phase 3, randomised controlled trial. Lancet Neurol. Aug 2022;21(8):693–703. doi:10.1016/s1474-4422(22)00209-5

4. Giovannoni G, Comi G, Cook S, et al. A placebo-controlled trial of oral cladribine for relapsing multiple sclerosis. N Engl J Med. Feb 4 2010;362(5):416–26. doi:10.1056/NEJMoa0902533

5. Hauser SL, Waubant E, Arnold DL, et al. B-cell depletion with rituximab in relapsing-remitting multiple sclerosis. N Engl J Med. Feb 14 2008;358(7):676–88. doi:10.1056/NEJMoa0706383

6. Hernán MA, Robins JM. Using Big Data to Emulate a Target Trial When a Randomized Trial Is Not Available. Am J Epidemiol. Apr 15 2016;183(8):758–64. doi:10.1093/aje/kwv254

7. König M, Lorentzen Å R, Torgauten HM, et al. Humoral immunity to SARS-CoV-2 mRNA vaccination in multiple sclerosis: the relevance of time since last rituximab infusion and first experience from sporadic revaccinations. J Neurol Neurosurg Psychiatry. Jan 2023;94(1):19–22. doi:10.1136/jnnp-2021-327612

8. König M, Torgauten HM, Tran TT, et al. Immunogenicity and Safety of a Third SARS-CoV-2 Vaccine Dose in Patients With Multiple Sclerosis and Weak Immune Response After COVID-19 Vaccination. JAMA Neurol. Mar 1 2022;79(3):307–309. doi:10.1001/jamaneurol.2021.5109

9. Moen SM, Harbo HF, Sowa P, Celius EG, Nygaard GO, Beyer MK. [MRI in multiple sclerosis]. Tidsskr Nor Laegeforen. Sep 2016;136(16):1373–6. MR-undersøkelser ved multippel sklerose. doi:10.4045/tidsskr.15.1361

10. Torgauten HM, Myhr KM, Wergeland S, Bo L, Aarseth JH, Torkildsen O. Safety and efficacy of rituximab as first- and second line treatment in multiple sclerosis - A cohort study. Mult Scler J Exp Transl Clin. Jan-Mar 2021;7(1):2055217320973049. doi:10.1177/2055217320973049

11. Thompson AJ, Banwell BL, Barkhof F, et al. Diagnosis of multiple sclerosis: 2017 revisions of the McDonald criteria. Lancet Neurol. Feb 2018;17(2):162–173. doi:10.1016/s1474-4422(17)30470-2

12. Hernán M, Robins J. Causal Inference: What If. 2020. Accessed June 14, 2024. https://www.hsph.harvard.edu/miguel-hernan/causal-inference-book/

13. Stensrud MJ, Aalen JM, Aalen OO, Valberg M. Limitations of hazard ratios in clinical trials. Eur Heart J. May 1 2019;40(17):1378–1383. doi:10.1093/eurheartj/ehy770

14. Gavoille A, Rollot F, Casey R, et al. Acute Clinical Events Identified as Relapses With Stable Magnetic Resonance Imaging in Multiple Sclerosis. JAMA Neurol. Jul 1 2024;doi:10.1001/jamaneurol.2024.1961

15. Comi G, Cook SD, Giovannoni G, et al. MRI outcomes with cladribine tablets for multiple sclerosis in the CLARITY study. J Neurol. Apr 2013;260(4):1136–46. doi:10.1007/s00415-012-6775-0

16. Comi G, Cook S, Rammohan K, et al. Long-term effects of cladribine tablets on MRI activity outcomes in patients with relapsing-remitting multiple sclerosis: the CLARITY Extension study. Ther Adv Neurol Disord. 2018;11:1756285617753365. doi:10.1177/1756285617753365

17. Hatcher SE, Waubant E, Nourbakhsh B, Crabtree-Hartman E, Graves JS. Rebound Syndrome in Patients With Multiple Sclerosis After Cessation of Fingolimod Treatment. JAMA Neurol. Jul 1 2016;73(7):790–4. doi:10.1001/jamaneurol.2016.0826

18. Pfeuffer S, Rolfes L, Hackert J, et al. Effectiveness and safety of cladribine in MS: Real-world experience from two tertiary centres. Mult Scler. Feb 2022;28(2):257–268. doi:10.1177/13524585211012227

19. Nygaard GO, Torgauten H, Skattebøl L, et al. Risk of fingolimod rebound after switching to cladribine or rituximab in multiple sclerosis. Mult Scler Relat Disord. Jun 2022;62:103812. doi:10.1016/j.msard.2022.103812

20. Roos I, Sharmin S, Malpas C, et al. Effectiveness of cladribine compared to fingolimod, natalizumab, ocrelizumab and alemtuzumab in relapsing-remitting multiple sclerosis. Mult Scler. Aug 1 2024:13524585241267211. doi:10.1177/13524585241267211

21. Spelman T, Frisell T, Piehl F, Hillert J. Comparative effectiveness of rituximab relative to IFN-β or glatiramer acetate in relapsing-remitting MS from the Swedish MS registry. Mult Scler. Jul 2018;24(8):1087–1095. doi:10.1177/1352458517713668

22. Meier S, Willemse EAJ, Schaedelin S, et al. Serum Glial Fibrillary Acidic Protein Compared With Neurofilament Light Chain as a Biomarker for Disease Progression in Multiple Sclerosis. JAMA Neurol. Mar 1 2023;80(3):287–297. doi:10.1001/jamaneurol.2022.5250

23. Benkert P, Maleska Maceski A, Schaedelin S, et al. Serum Glial Fibrillary Acidic Protein and Neurofilament Light Chain Levels Reflect Different Mechanisms of Disease Progression under B-Cell Depleting Treatment in Multiple Sclerosis. Ann Neurol. Oct 16 2024;doi:10.1002/ana.27096

24. Salter A, Fox RJ, Newsome SD, et al. Outcomes and Risk Factors Associated With SARS-CoV-2 Infection in a North American Registry of Patients With Multiple Sclerosis. JAMA Neurol. Jun 1 2021;78(6):699–708. doi:10.1001/jamaneurol.2021.0688

25. Sormani MP, De Rossi N, Schiavetti I, et al. Disease-Modifying Therapies and Coronavirus Disease 2019 Severity in Multiple Sclerosis. Ann Neurol. Apr 2021;89(4):780–789. doi:10.1002/ana.26028

26. Simpson-Yap S, De Brouwer E, Kalincik T, et al. Associations of Disease-Modifying Therapies With COVID-19 Severity in Multiple Sclerosis. Neurology. Nov 9 2021;97(19):e1870–e1885. doi:10.1212/wnl.0000000000012753

27. Torgauten HM, Onyango TB, Ljostveit S, et al. Hospitalisations and humoral COVID-19 vaccine response in vaccinated rituximab-treated multiple sclerosis patients. Mult Scler Relat Disord. Jul 15 2024;89:105770. doi:10.1016/j.msard.2024.105770

28. Langer-Gould AM, Smith JB, Gonzales EG, Piehl F, Li BH. Multiple Sclerosis, Disease-Modifying Therapies, and Infections. Neurol Neuroimmunol Neuroinflamm. Nov 2023;10(6)doi:10.1212/nxi.0000000000200164

29. Hauser SL, Bar-Or A, Comi G, et al. Ocrelizumab versus Interferon Beta-1a in Relapsing Multiple Sclerosis. N Engl J Med. Jan 19 2017;376(3):221–234. doi:10.1056/NEJMoa1601277

30. Steinman L, Fox E, Hartung HP, et al. Ublituximab versus Teriflunomide in Relapsing Multiple Sclerosis. N Engl J Med. Aug 25 2022;387(8):704–714. doi:10.1056/NEJMoa2201904

31. Hauser SL, Bar-Or A, Cohen JA, et al. Ofatumumab versus Teriflunomide in Multiple Sclerosis. N Engl J Med. Aug 6 2020;383(6):546–557. doi:10.1056/NEJMoa1917246

32. Luna G, Alping P, Burman J, et al. Infection Risks Among Patients With Multiple Sclerosis Treated With Fingolimod, Natalizumab, Rituximab, and Injectable Therapies. JAMA Neurol. Feb 1 2020;77(2):184–191. doi:10.1001/jamaneurol.2019.3365

